# An assessment of interobserver agreement on lesion size, morphology and clinical phenotype in cutaneous leishmaniasis caused by *Leishmania aethiopica* in Ethiopia

**DOI:** 10.1101/2024.12.09.24318700

**Authors:** Amel Beshir Mohammed, Fewzia Shikur Mohammed, Feleke Tilahun Zewdu, Shimelis Doni Nigusse, Saba Lambert, Michael Marks, Stephen L. Walker, Endalamaw Gadisa, SHARP collaboration

**Affiliations:** ALERT Hospital, Addis Ababa, Ethiopia; Department of Dermatovenereology, College of Health Sciences, Addis Ababa University, Addis Ababa, Ethiopia; Boru Meda Hospital, Boru Meda, Ethiopia; Department of Epidemiology and Biostatistics, Institute of Public Health, College of Medicine and Health Sciences, University of Gondar, Gondar, Ethiopia; Department of Malaria and Neglected Tropical Disease Research, Armauer Hansen Research Institute, Addis Ababa, Ethiopia; Department of Paediatrics and Neonatal Health Nursing, College of Health Sciences, Debre Tabor University, Debre Tabor, Ethiopia; Department of Infection Biology, Faculty of Infectious and Tropical Diseases, London School of Hygiene & Tropical Medicine, London, UK; Department of Global Health and Development, London School of Hygiene & Tropical Medicine, London, UK; School of Public Health, Addis Ababa University, Addis Ababa, Ethiopia; Department of Medical Statistics, Faculty of Epidemiology and Population Health, London School of Hygiene & Tropical Medicine, London, UK; Department of Clinical Research, Faculty of Infectious and Tropical Diseases, London School of Hygiene & Tropical Medicine, London, UK; Hospital for Tropical Diseases, University College London Hospitals NHS Foundation Trust, London, UK; University College London, London, UK

## Abstract

**Introduction:** Cutaneous leishmaniasis (CL) remains a major public health challenge, especially in endemic regions like Ethiopia, where an estimated 40,000 new cases occur annually. Effective treatment evaluation for CL relies on consistent clinical assessments, yet variability in lesion descriptions can complicate reliable outcome measures.

**Methods:** We conducted an inter-reliability study of clinicians’ evaluations of CL lesion morphology and size at ALERT Hospital, Addis Ababa. Twelve clinicians independently examined 12 patients with parasitologically confirmed CL, each clinician assessing lesion morphology, size, and severity.

**Results:** We found high consistency in reporting major morphological categories (e.g., plaques) but significant variability in secondary features like dyspigmentation and scale, as well as mucosal involvement. Lesion size measurements showed limited variability, suggesting its reliability as a potential measure for future clinical trials. Disparities in severity assessments highlight the need for a standardized scoring system in CL.

**Discussion:** Our findings underscore the importance of training for consistent, high-quality clinical evaluations of CL and suggests that lesion size could be a reproducible outcome measure in treatment efficacy trials.

## Introduction

Cutaneous leishmaniasis (CL) is the most common form of leishmaniasis. The disease causes skin lesions on exposed parts of the body at the site of an infected sandfly bite. With or without treatment CL lesions can lead to life-long scars and result in serious disability or stigma. It is estimated that 600 000 to 1 million new cases occur worldwide annually with 40,000 new cases of CL in Ethiopia ^1^. Three *Leishmania* species have been reported in Ethiopia, but the vast majority of cases are thought to be due to *L aethiopica*.

*L. aethiopica* is associated with a variety of clinical phenotypes. The most frequent presentation is localized CL (LCL), but infection is also associated with mucocutaneous leishmaniasis (MCL), and diffuse CL (DCL). LCL usually starts as a papule that gradually enlarges forming nodules or plaques. Ulceration may occur with *L. aethiopica*. MCL can occur either via the bite of the sand fly at the border of mucosal surfaces or through extension from an adjacent cutaneous lesion. It most commonly involves the nasal mucosa but the lips and oral mucosa can also be involved. DCL is a chronic and progressive disorder which starts with papular or nodular lesions which become larger and more numerous^2^.

Despite the large number of affected individuals, studies of the clinical manifestations of *L. aethiopica*, including the morphology of the lesions, progression of the disease and response to therapy, are limited. A study in North-west Ethiopia reported the most common features to be induration, erythema, ulceration, and crusting.

The WHO strategic framework for integrated control and management of “skin-related” NTDs highlighted anti-microbial therapy for CL as a research gap and this is particularly the case for treatment of *L. aethiopica* infection^3,4^. Robust outcome measures are required for treatment efficacy studies. Outcome measures in CL in studies are based on clinical evaluation of individual lesions including re-epithelization in ulcerated lesions and flattening in non-ulcerated ones and lesion measurement. In 2018, a proposal for harmonized clinical trial methodologies for cutaneous leishmaniasis was published to develop a consensus and standardize clinical evaluation as a measure of disease response and cure in CL^5^.

Given the reliance on clinical observations of lesion morphology and severity it is therefore essential to evaluate whether the clinical evaluation of individual physicians is comparable. We therefore conducted a study to assess the inter-rater reliability of clinicians’ assessment of lesion morphology and size in individuals affected by CL in Ethiopia.

## Methods

The study was conducted nested within a larger cohort study of CL in Ethiopia. As part of this larger study, we conducted an inter-observer exercise at ALERT comprehensive specialized Hospital, Addis Ababa^6^.

Examiners were selected from the clinical staff of ALERT Hospital and members of the Skin Health Africa Research Programme (SHARP) team. We collected information on their previous training in skin disease and experience of managing CL.

Clinicians at ALERT Hospital identified 12 individuals with a parasitologically confirmed diagnosis of CL who were invited to participate in the study. A standardized proforma was used to capture routine clinical data about each affected individual. For individuals with more than one clinical lesion of CL, the treating clinician indicated the “index” lesion to be assessed in the inter-observer assessment exercise. This “index” lesion was the lesion identified by the affected individual as the one of greatest concern prior to the start of the exercise.

A computer generated latin-square was used to randomize an order of evaluation for each observer and affected individual. Each observer was assigned a unique letter and each affected individual a unique number. To measure inter-observer agreement each observer independently examined all 12 individuals with CL once for a period of 5 minutes.

Examiners recorded on a standard data collection sheet their assessment of each “index” lesion. Standardized definitions of CL developed for our cohort study (Box 1) were used^6^. For each lesion examiners were asked to record morphology, size and the presence or absence of clinical features including induration, ulceration, crust, hyper/hypopigmentation, scarring and mucosal involvement. Examiners measured the largest diameter of the lesion using a disposable tape measure. Examiners were asked to provide an assessment of the clinical phenotype of CL and to classify the disease severity as mild or moderate or severe. All data were double entered into an electronic study database for analysis.

As there is no published data on the degree of agreement in assessing cases of Cutaneous Leishmaniasis we used a convenience sample size based on ensuring we had a range of both examiners and patients with clinical differing clinical phenotypes. The primary planned analysis was descriptive. We report the proportion of affected individuals judged to have each feature by the examiners. For lesion size we describe variability by reporting the median and IQR lesion size reported for each index lesion. Analysis was conducted in R version 4.2.1.

Ethical approval was obtained from AHRI/ALERT Ethics Review Committee (Ref: PO/23/21), the National Research Ethics Review Committee (Ref: 7/2-506/m259/35), and the London School of Hygiene and Tropical Medicine (Ref: 26421). Written individual informed consent was obtained from all participants (examiners and affected individuals) in the study.

## Results

Twelve affected individuals agreed to participate in the study. Seven were male and five female. The median age was 25 years. Based on the treating clinician’s assessment six had been diagnosed with LCL, four with MCL and two with mucosal CL. All twelve individuals were currently receiving inpatient treatment with intramuscular sodium stibogluconate.

Twelve examiners took part in the study. Seven of the clinicians were male and 5 were female. The median age of examiners was 39. The majority (n = 10, 83%) were clinicians with specialist postgraduate training in dermatology and six (50%) were accredited dermatologists. (Table 1).

**Table 1:** Characteristics of individuals conducting the examinations.

|  |  |
| --- | --- |
| Median Age (years) | 39 |
| Male | 7 |
| Years experience | 8.3 (SD 6.8) |
| Clinical specialty |  |
| Dermatologist | 6 |
| Dermatology Trainee | 3 |
| Infectious Diseases | 1 |
| MSc in Dermatology | 1 |
| GP with Dermatology postgraduate diploma | 1 |

Some clinical features appeared to be more consistently reported by examiners than others (Table 2). For lesion morphology all examiners agreed on the presence of plaques in one individual, whilst 10 or more examiners agreed on the presence of plaques in five. In the remaining individuals with CL either eight or nine examiners assessed the lesion to be a plaque. All examiners agreed there were no nodular lesions in eight individuals. For the remaining four affected individuals one to four examiners felt there were nodular lesions. There was greater variability in reporting of the presence or absence of papules. For five individuals all examiners agreed papules were absent whereas for seven individuals between one to five examiners assessed they were present.

**Table 2:** The number of times each clinical feature was reported for each patient.

| Affected Individual ID | Treating Clinician Phenotype | Erythema | Indurated | Ulcerated | Crusted | Scaly | Scar | Hypopigmented | Hyperpigmented | Papule | Nodule | Plaque | Mucosa |
| --- | --- | --- | --- | --- | --- | --- | --- | --- | --- | --- | --- | --- | --- |
| 1 | LCL | 3 | 12 | 2 | 11 | 5 | 4 | 1 | 5 | 5 | 0 | 11 | 0 |
| 2 | LCL | 7 | 11 | 0 | 12 | 11 | 0 | 1 | 10 | 0 | 2 | 8 | 7 |
| 3 | Pure Mucosal | 11 | 12 | 1 | 1 | 10 | 0 | 1 | 0 | 1 | 1 | 8 | 12 |
| 4 | MCL | 12 | 11 | 9 | 2 | 6 | 1 | 0 | 2 | 1 | 0 | 9 | 11 |
| 5 | MCL | 12 | 12 | 0 | 8 | 10 | 1 | 5 | 5 | 0 | 0 | 10 | 11 |
| 6 | MCL | 12 | 12 | 4 | 7 | 1 | 6 | 5 | 8 | 2 | 0 | 11 | 12 |
| 7 | LCL | 6 | 12 | 0 | 0 | 2 | 11 | 0 | 10 | 5 | 0 | 11 | 2 |
| 8 | LCL | 5 | 11 | 0 | 2 | 10 | 2 | 1 | 8 | 0 | 0 | 12 | 0 |
| 9 | LCL | 10 | 9 | 0 | 3 | 11 | 0 | 1 | 4 | 2 | 0 | 10 | 1 |
| 10 | Pure Mucosal | 11 | 9 | 1 | 0 | 9 | 1 | 3 | 1 | 0 | 0 | 9 | 12 |
| 11 | LCL | 8 | 9 | 2 | 12 | 2 | 0 | 0 | 5 | 0 | 3 | 9 | 0 |
| 12 | MCL | 11 | 11 | 0 | 10 | 10 | 2 | 0 | 8 | 3 | 1 | 9 | 9 |

Involvement of the mucosa was reported by all examiners for three individuals and by 11 examiners in two others. For three individuals with CL all examiners agreed there was no mucosal involvement and for a further two individuals only one or two examiners assessed the mucosa to be affected. For two individuals there was more marked disagreement with seven and nine examiners reporting involvement and five and three examiners reporting no mucosal involvement.

For lesion features all examiners agreed on the presence of erythema in three individuals and at least 10 examiners agreed it was present in a further four. For the remaining individuals with CL there was marked variability with between three to eight examiners reporting erythema. For five individuals all examiners reported the presence of induration and for a further four at least 11 examiners felt induration was present. For the remaining three affected individuals nine examiners felt induration was present and three felt it was absent.

For six individuals with CL all examiners agreed that there was no evidence of ulceration. For the remaining six patients the number of examiners reporting ulceration varied from one to nine. For two individuals all 12 examiners agreed that there was evidence of crust and for two individuals all examiners agreed crust was absent. For the remaining eight individuals with CL the number of examiners reporting crust varied from one to eleven.

There were no individuals with CL whom examiners were in agreement on the presence or absence of scale but there were six where at least 10 examiners agreed scale was present and three where at least 10 examiners agreed it was absent.

Finally, all examiners agreed there was no evidence of scarring in four individuals. For the remaining eight between one and eleven examiners deemed scarring to be present.

Estimates of the reported maximum lesion diameter were broadly consistent across examiners but graphical visualization with a modified Bland-Altman plot suggests variation increased with the size of the lesion (Figure 1)^7^.

**Figure 1:**
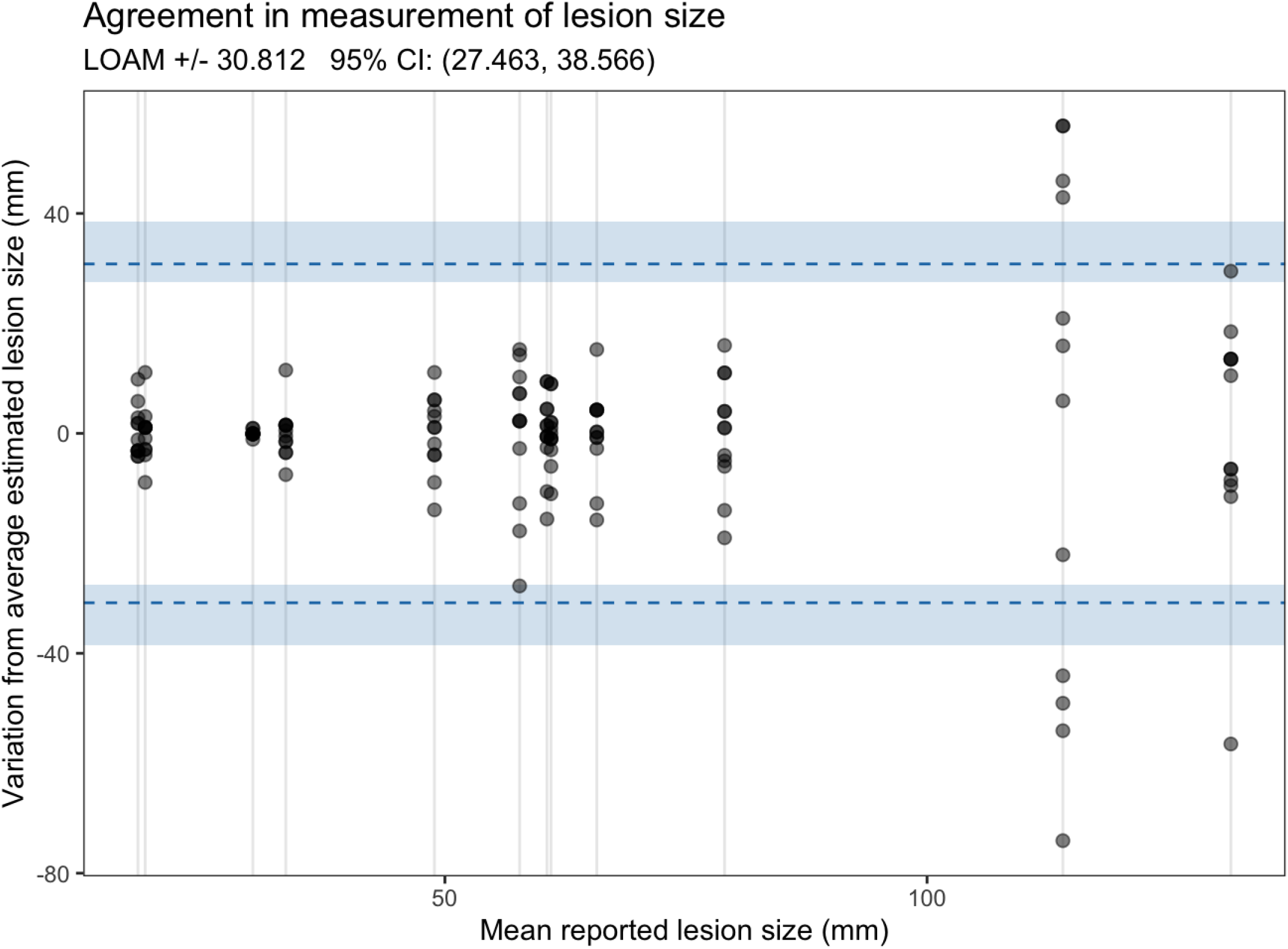
Bland-Altman plot showing variation in assessment of lesion size.

The majority of observers assessed that two third of patients had moderate to severe disease with the remaining affected individuals assessed as having mild-moderate disease. There were only two individuals with CL in whom there was a 2-grade discrepancy in the assessment of severity but there were no individuals in whom all examiners assigned the same severity score and only one in whom there was more than 80% agreement. Disagreement about mucosal involvement also meant that there were only four (33%) individuals with CL where all 12 examiners and the treating clinicians agreed on the clinical phenotype (Table 3).

**Table 3:** Clinical Phenotype and Severity as judged by the examiners.

| Patient | Treating Clinician Phenotype | Phenotype Assessment |  |  |  | Severity Assessment |  |  |
| --- | --- | --- | --- | --- | --- | --- | --- | --- |
|  |  | DCL | LCL | MCL | Pure Mucosal | Mild | Moderate | Severe |
| 1 | LCL | 0 | 12 | 0 | 0 | 0 | 8 | 4 |
| 2 | LCL | 0 | 5 | 7 | 0 | 1 | 7 | 4 |
| 3 | Pure Mucosal | 0 | 0 | 5 | 7 | 0 | 5 | 7 |
| 4 | MCL | 0 | 2 | 10 | 0 | 0 | 7 | 5 |
| 5 | MCL | 0 | 2 | 10 | 0 | 0 | 2 | 10 |
| 6 | MCL | 0 | 0 | 12 | 0 | 0 | 3 | 9 |
| 7 | LCL | 1 | 9 | 2 | 0 | 0 | 5 | 7 |
| 8 | LCL | 0 | 12 | 0 | 0 | 7 | 4 | 1 |
| 9 | LCL | 0 | 11 | 1 | 0 | 5 | 6 | 1 |
| 10 | Pure Mucosal | 0 | 0 | 10 | 2 | 6 | 3 | 3 |
| 11 | LCL | 0 | 12 | 0 | 0 | 9 | 3 | 0 |
| 12 | MCL | 0 | 3 | 9 | 0 | 1 | 4 | 7 |

## Discussion

In the first ever study evaluating the inter-observer reliability of clinical examination to assess lesion morphology in CL we demonstrated considerable variation in the reporting of several key clinical features. Reporting of overall lesion morphology as plaque, nodule or papule was generally highly consistent but the presence or absence of secondary clinical characteristics such as dyspigmentation or the presence of scale were more variable.

Lesion size is proposed as a key characteristic for assessing treatment outcomes in future trials^5^. Reassuringly, we showed that there was limited variation in the assessed size of lesions when measured by multiple independent assessors, although there seemed to be evidence that variability was highest for larger CL lesions. Overall, these data suggest that lesion size assessment is likely to be a reliable measure in both observational and interventional prospective studies.

We found assessment of mucosal involvement to be variable. As this has important implications for treatment and characterization for clinical trials it may be advisable in some cases to obtain an assessment by an ear, nose and throat specialist to allow accurate characterization. There is not yet a standardized definition of severity for CL^6^. In the absence of such a system we found little consensus amongst clinicians when assessing individuals with CL which poses challenges in reliable reporting of their characteristics in both observational and interventional studies. Development of a validated severity score should be considered to help standardize practice.

Inter-observer variability is well recognized in clinical assessment particularly when assessing subjective features of disease. Similar to our findings, previous studies in other NTDs have shown that the degree of reliability may vary for different clinical features within a given disease. For example, diagnosis of scabies by healthcare workers has been shown to vary markedly depending on the severity of the infestation^8,9^. Similarly for leprosy studies have shown marked differences in inter-observer reliability in determining the presence or absence of nerve thickening or in the reliability of different methods of assessing sensory loss within skin lesions^10^.

The new WHO roadmap for NTDs focuses on strengthening responses to skin-NTDs^3^. Achieving this will require considerable investment in training of healthcare workers in the diagnosis and management of a range of skin diseases including CL. Given the degree of variability between expert examiners in this study our data suggest such efforts will require considerable effort to ensure high-quality, reliable and reproducible clinical diagnosis and assessment of clinical healing parameters.

Our study has limitations. Firstly, all affected individuals were inpatients receiving care for CL at a referral centre. This is likely to have resulted in selection of individuals with more severe disease than might be seen elsewhere. Similarly, all individuals were receiving treatment and therefore their lesions might differ from untreated ones. Whilst this might affect lesion morphology, we believe it is unlikely to have affected our assessment of inter-examiner reliability. The generalizability of our findings to regions of the world where other *Leishmania* species are endemic is unclear, particularly in settings where ulcerated lesions may be more common.

Assessment of inter-examiner reliability for multiple examiners is challenging. Conventionally kappa-scores are calculated to compare the performance of an examiner against a reference-standard. Multi-examiner weighted kappa scores have been developed to extend this approach to situations broadly analogous to our study design. However, both conventional and multi-examiner kappa scores have weaknesses^11^. Disagreements limited to a small number of individuals being examined can have an outsize impact on kappa scores even when there is near universal consensus on the majority individuals. We therefore opted to only present descriptive statistics. Finally, our study focuses on the assessment of individuals with known CL by experts. Further studies are needed to assess the performance of other cadres of healthcare workers in the initial evaluation of individuals with suspected CL.

Trials of novel and improved therapeutics for CL are urgently needed. To provide comprehensive data on efficacy such trials will need to combine reliable and reproducible parasitological, clinical and patient reported outcome measures. Measurement of lesion size is likely to be a critical component of such measures, and our data are reassuring that this can be reliably measured by multiple independent assessors. More work is needed to identify and improve assessment of other clinical features in order to develop robust trial outcome measures for CL.

## Data Availability

All data is contained within the paper

## Acknowledgements

We wish to thank the individuals and communities for their participation in the work of the Skin Health Africa Research Programme.

## BOX 1

Operational definitions of Cutaneous Leishmaniasis (CL)

- CL is diagnosed in a person with skin and/or mucosal lesions with evidence of *Leishmania* infection in the affected tissues characterised by the presence of *Leishmania* amastigotes on smear microscopy or growth of *Leishmania* promastigotes in culture or the detection of *Leishmania* DNA by polymerase chain reaction.
- Localized CL: A confirmed case of leishmaniasis, with no mucosal involvement, characterized by ten or fewer cutaneous papules and/or nodules and/or plaques with or without ulceration involving one body site
- Multi-regional localized CL: A confirmed case of leishmaniasis, with no mucosal involvement, characterized by ten or fewer cutaneous papules and/or nodules and/or plaques with or without ulceration involving two or more body sites
- Mucocutaneous leishmaniasis: A confirmed case of leishmaniasis characterized by ten or fewer papules and/or nodules and/or plaques with or without ulceration involving skin and an adjacent mucosal surface
- Mucosal leishmaniasis: A confirmed case of leishmaniasis characterized by papules and/or nodules and/or plaques with or without ulceration involving exclusively a mucosal surface
- Diffuse CL: A confirmed case of leishmaniasis characterized by eleven or more papules and/or nodules and/or plaques with or without mucosal involvement.

Body sites are classified as:

1. Head and neck
2. Torso - anterior (including genitalia)
3. Torso - posterior (including buttocks)
4. Right upper limb
5. Left upper limb
6. Right lower limb
7. Left lower limb

## References

1 Alvar J, Vélez ID, Bern C, et al. Leishmaniasis worldwide and global estimates of its incidence. PLoS ONE. 2012; 7: 35671.

2 Bryceson AD. Diffuse cutaneous leishmaniasis in Ethiopia. I. The clinical and histological features of the disease. Trans R Soc Trop Med Hyg 1969; 63: 708–37.

3 Ending the neglect to attain the sustainable development goals: a strategic framework for integrated control and management of skin-related neglected tropical diseases. https://www.who.int/publications/i/item/9789240051423 (accessed June 25, 2024).

4 Griensven J van, Gadisa E, Aseffa A, Hailu A, Beshah AM, Diro E. Treatment of Cutaneous Leishmaniasis Caused by Leishmania aethiopica: A Systematic Review. PLOS Neglected Tropical Diseases 2016; 10: e0004495.

5 Olliaro P, Grogl M, Boni M, et al. Harmonized clinical trial methodologies for localized cutaneous leishmaniasis and potential for extensive network with capacities for clinical evaluation. PLOS Neglected Tropical Diseases 2018; 12: e0006141.

6 Mohammed AB, Mohammed FS, Zewdu FT, et al. Protocol for a prospective observational cohort study of cutaneous leishmaniasis in Ethiopia. NIHR Open Res 2023; 3: 49.

7 Christensen HS, Borgbjerg J, Børty L, Bøgsted M. On Jones et al.’s method for extending Bland-Altman plots to limits of agreement with the mean for multiple observers. BMC Medical Research Methodology 2020; 20: 304.

8 Walker SL, Collinson S, Timothy J, et al. A community-based validation of the International Alliance for the Control of Scabies Consensus Criteria by expert and non-expert examiners in Liberia. PLOS Neglected Tropical Diseases 2020; 14: e0008717.

9 Osti MH, Sokana O, Lake S, et al. The body distribution of scabies skin lesions. JEADV Clinical Practice; n/a. DOI:10.1002/jvc2.26.

10 Ponnighaus JM, Fine PE, Bliss L. Certainty levels in the diagnosis of leprosy. Int J Lepr 1987; 55: 454–62.

11 Landis JR, Koch GG. The measurement of observer agreement for categorical data. Biometrics 1977; 33: 159–74.

